# Re-analysis of genetic risks for Chronic Fatigue Syndrome from 23andMe data finds few remain

**DOI:** 10.1101/2020.10.27.20220939

**Authors:** Felice L. Bedford, Bastian Greshake Tzovaras

## Abstract

It is tempting to mine the abundance of DNA data that is now available from direct-to-consumer genetic tests but this approach also has its pitfalls A recent study put forth a list of 50 single nucleotide polymorphisms (SNPs) that predispose to Chronic Fatigue Syndrome (CFS), a potentially major advance in understanding this still mysterious condition. However, only the patient cohort data came from a commercial company (23andMe) while the control was from a genetic database. The extent to which 23andMe data agree with genetic reference databases is unknown. We reanalyzed the 50 purported CFS SNPs by comparing to control data specifically from 23andMe which are available through public platform OpenSNP. In addition, large high-quality database ALFA was used as an additional control. The analysis lead to dramatic change with the top of the leaderboard for CFS risk reduced and reversed from an astronomical 129,000 times to 0.8. Errors were found both within 23andMe data and the original study-reported Kaviar database control. Only 3 of 50 SNPs survived initial study criterion of at least twice as prevalent in patients, EFCAB4B, involved in calcium ion channel activation, LINC01171, and MORN2 genes. We conclude that the reported top-50 deleterious polymorphisms for Chronic Fatigue Syndrome were more likely the top-50 errors in the 23andMe and Kaviar databases. In general, however, correlation of 23andMe control with ALFA was a respectable 0.93, suggesting an overall usefulness of 23andMe results for research purposes but only if caution is taken with chips and SNPs.

## 1 Introduction

As part of a growing number of researchers that advocate using the plentiful genetic data from direct to consumer testing (1), we are also aware of its pitfalls. Recently, a study in Frontiers in Pediatrics (2) described an ambitious project to elucidate genetic predispositions for Chronic Fatigue Syndrome (CFS), a not-uncommon condition of debilitating fatigue, immune dysregulation, and central nervous system impairment. The study analyzed the approximate 500,000 genetic single nuclear polymorphisms (snps) resulted by the commercial 23andMe genetic testing company in people with CFS.

Two items from the Perez et al. results were immediate red flags. They showed the top 50 deleterious snps with the greatest difference in frequency between CFS patients and control data. At the very top of the leaderboard was a snp on the gene GPBAR1 that was 129,000 times more prevalent in CFS patients. If this finding is accurate then the authors may well have discovered THE genetic cause of CFS rather than just a predisposition. In addition, the CYP2D6 gene on their list is recognizable as one that in the past, for a different snp, had the majority of 23andMe customers believing they had a poor xenobiotic metabolizer phenotype that only actually affects less than one percent of the population (from Snpedia 3).

In the Perez et al. study, the data used to compare to the 23andMe CFS participants did not come from 23andMe. Instead, they relied on published frequencies in the Kaviar database, a compilation from multiple projects (4). Unless the control data comes from the identical source as the experimental data, any differences in quality or population constituency between the two datasets may lead to inaccurate conclusions when compared. How many of the reported frequency ratios between CFS patients and controls would remain noteworthy if both data come from the same source? The study established a criterion that a snp should be at least twice as prevalent in CFS patients, a ratio of 2, to be of note. In addition, how closely do 23andMe data match published genetic reference data? Errors in direct-to-consumer genetic tests have been reported (5).

To address these questions, we accessed a publicly available control set of genomes from 23andMe participants. We reanalyzed the frequency ratio of the highly prevalent snps in CFS from the Perez et al. study to this more appropriate control. We also incorporated an additional new large high-quality online control dataset for further comparison among control datasets.

## 2 Method

To provide control data from 23andMe for the 50 top-predisposing CFS snps, we accessed publicly available genome files on openSNP^1^, a platform co-founded and maintained by one of us (BGT). OpenSNP allows individuals to upload their own genetic results from a variety of commercial test companies for public sharing (6). The allele frequencies and genotypes for these SNPs were calculated for all 23andMe data sets present in openSNP on 2020-06-19. While self-selection of participants can skew a dataset, the platform also allows phenotypes of interest to be added by participants. We noted the absence of CFS and Myalgic Encephalomyelitis (ME) on the list of phenotypes which provided an initial confidence that the dataset does not contain an overrepresentation of CFS patients compared to the general population.

For the additional control: The allele frequency aggregation project (ALFA) project was developed as part of the National Center for Biotechnology Information (NCBI) database of genotypes and phenotypes (dbGAP) and had their inaugural release on March 10, 2020 (7). It contains a high-quality aggregate of over 1200 studies with a goal of one million dbGAP subjects. ALFA (build 154, release date April 21, 2020) was accessed through NCBI dbSNP^2^. ALFA Europe was selected when available to match the population of 23andMe, primarily Americans of European decent. When unavailable, ALFA Global was the second choice, followed by GnomAD – exome and then 1000 Genome project if absent from ALFA entirely. When ALFA is referred to subsequently, it is a shortcut notation for the totality of this procedure.

To recalculate the ratio with the new controls, for the CFS data we used the frequencies provided by Perez et al. in Table 1. Since neither the table nor supplementary materials explicitly listed the alleles, we assumed that the frequencies always referred to the derived allele. In the event of multiple derived alleles at a position, we further assumed the most prevalent one was used. Spot checks of their Kaviar control supported that this was the study’s intended listing. The new ratios were recalculated for the 23andMe control and for the ALFA control and subsequently compared to the original ratios. The three control datasets at the 50 snps (Kaviar, 23andMe, ALFA) were compared to each other.

**Table 1.**
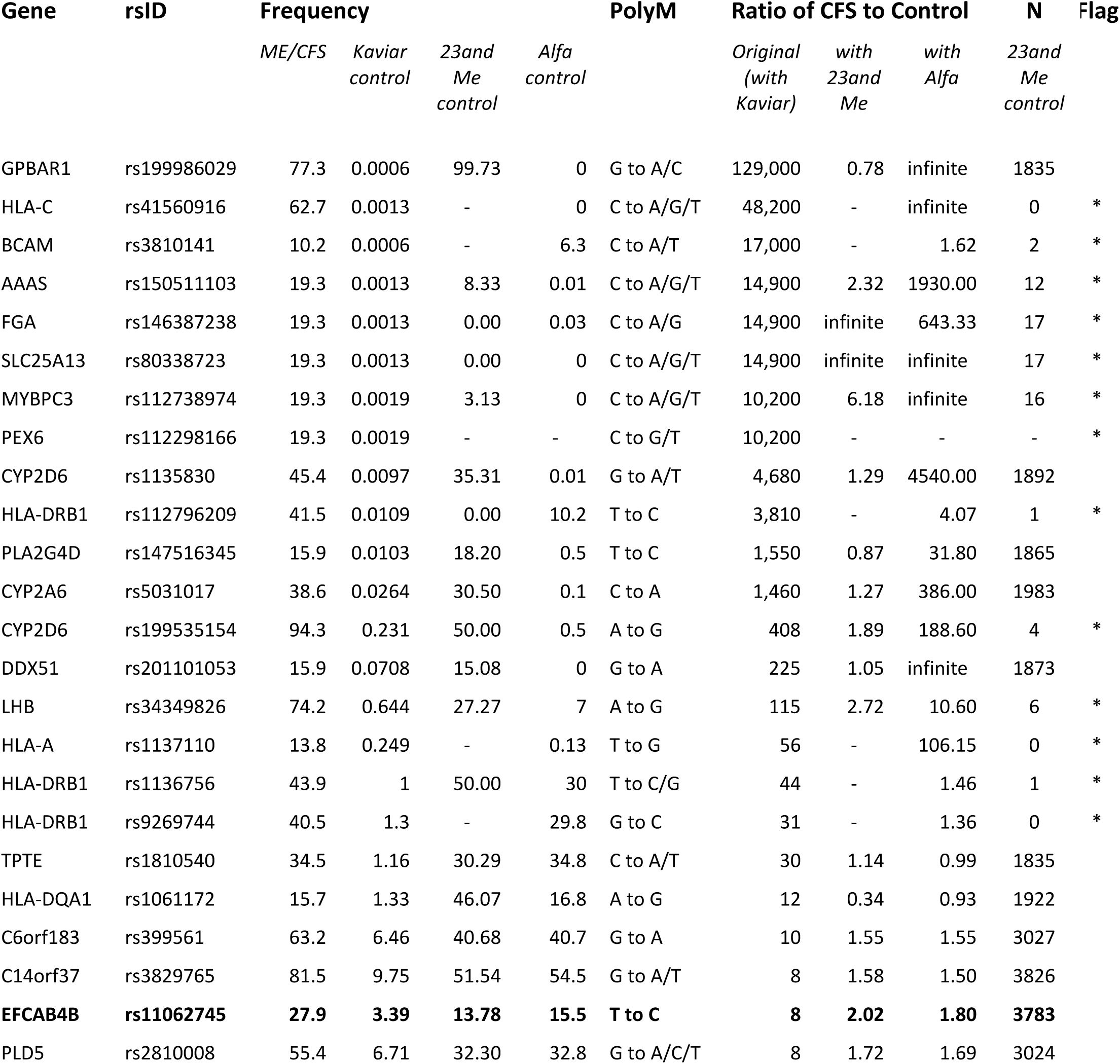
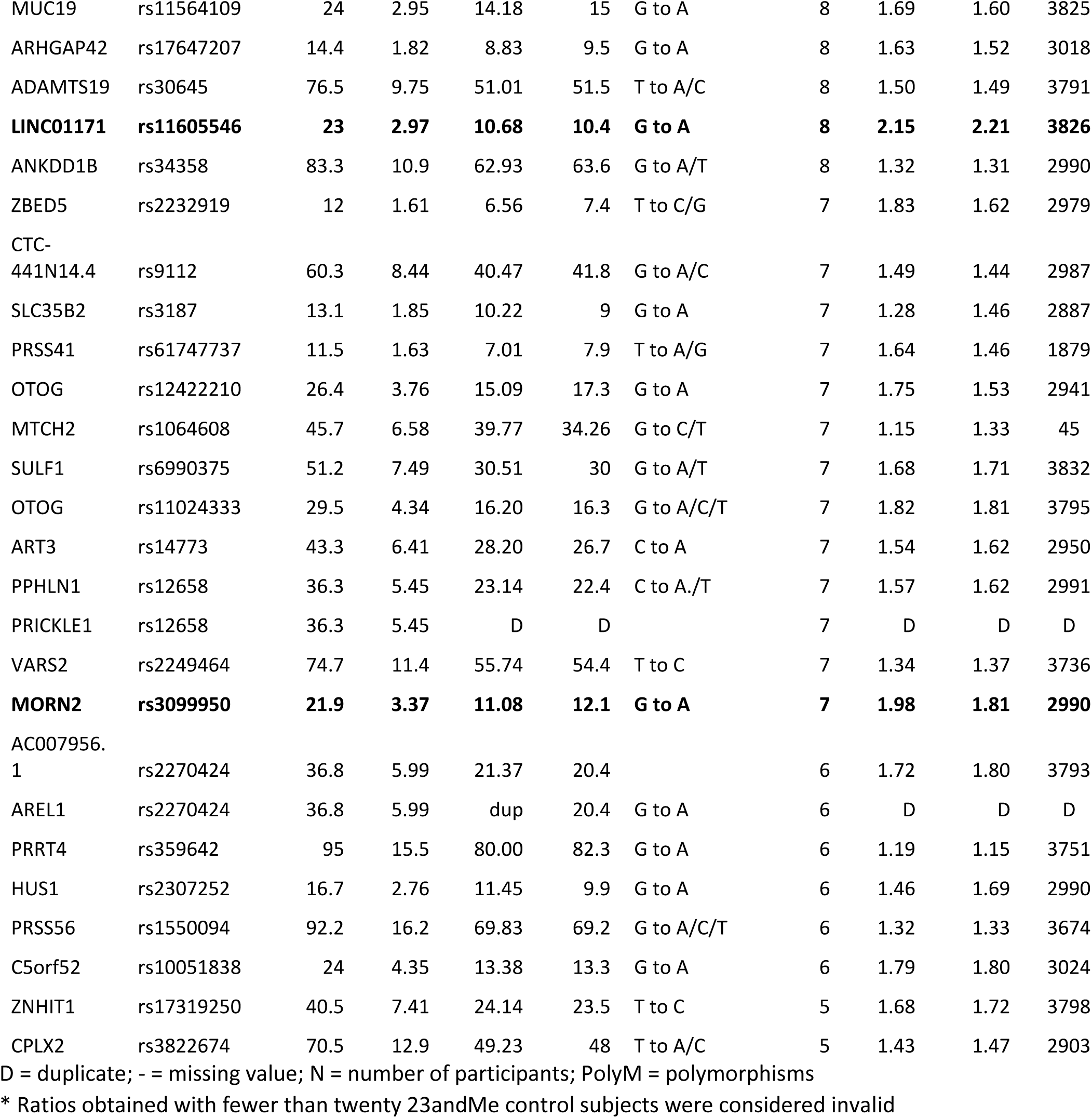
Recalculated ratios of polymorphism frequencies of CFS patients to controls.

## 3 Results

The recalculated ratio of allele frequency in CFS patients to control subjects using 23andMe control data, or where unavailable, ALFA frequencies, along with the elimination of 2 duplicates, found that only 11 of the 50 polymorphisms now exceeded a ratio of 2. That is, only 22 percent of the originally reported polymorphisms remained at the original study criterion that notable snps were at least double the frequency in CFS patients compared to unaffected controls.

Of the 11 remaining polymorphisms with a ratio that met the criterion, the majority, 7, could be based only on ALFA frequencies; and 1 was not reported on ALFA either. These 8 snps were only present in the 23andMe control data set in very few samples, ranging from 17 to 0, in contrast to a median of nearly 3000 samples in the 23andMe control data overall. These further 8 snps therefore were also not shown to have a higher prevalence in 23andMe CFS patients than in 23andMe controls. Dates of upload to openSNP hint that the early days of the 23andMe v5 chip could be a source of error. All 8 of these come from the top of the original ratio leaderboard (Table 1).

Only 3 snps of 50 remained, on genes EFCAB4B, LINC01171, and MORN2, that could be shown to meet the original criterion of at least double in CFS patients compared to a comparable 23andMe control, all hovering at a ratio of about 2.0. None of the astronomically high ratios of patients to controls could be shown to remain.

Table 1 presents the ratios of allele frequency in CFS patients to control subjects (original, recalculated with the new 23andMe control, recalculated with ALFA), the frequency of allele occurrence (original CFS patients, original Kaviar, new 23andMe control, new ALFA control), and the number of samples (23andMe control) for each of the snps reported in the original table of the study. Genotype frequencies found in the 23andMe control samples for each snp is in Supplementary Figure 1.

Comparison of the control datasets (Kaviar, 23andMe, ALFA) found 2 primary patterns. Most prevalent, 29 out of the 48 of the 23andMe control frequencies were in good agreement with ALFA with both being substantially higher than the reported Kaviar values, suggesting a Kaviar-related error. We call this error Type A. For 8 of the 48 snps, the 23andMe control frequencies were instead different (higher) than both ALFA and Kaviar, which were in good agreement with each other and point to a 23andMe error (Type B error). Like the missing 23andMe snps, Type B errors came from the top of the leaderboard and further accounts for the original astronomic reported ratios.

Concerning the two-red flag genes mentioned at the outset, both are Type B errors. The GPBAR1 snp for derived allele A was found in 97% of the 23andMe control sample compared to practically 0 in the other control data leading to a recalculation of the ratio of prevalence between CFS patients and controls from the reported 129,000 to 0.8, a reversal. Based on upload dates, this erroneous base A call may be traceable specifically to the v4 chip but would require exact test dates for confirmation. The CYP2D6 had 2 snps, one of which had more than a third erroneous 23andMe call for base A.

The correlation between the 23andMe control and ALFA control frequencies without the genes above. and without any 23andMe data having fewer than 50 participants, was a respectable 0.93.

## Discussion

The genetic predispositions reported for Chronic Fatigue Syndrome are not supported when reanalyzed with more appropriate control data including those drawn from the same 23andMe pool as the CFS patients. Out of the original 50 genomic positions presented to have the most prevalent deleterious polymorphisms among CFS sufferers, only three remained that met the original study criterion of at least twice as frequent as healthy individuals. The top-ranked risk factor on gene GPBAR1 with an astronomical ratio of 129,000 was reduced to the more sensible 0.8, which would, if anything, be a protective snp against CFS.

The erroneous odds ratios were found to originate from a mixture of errors in both 23andMe and in the reported Kaviar control dataset. The more dramatic frequencies that had been listed as dozens, hundreds, and thousands of times higher in CFS patients were due to seeming 23andMe peculiarities of very high frequencies for minor alleles. We found these 23andMe errors to either also be present in high numbers in the 23andMe controls (Genes GPBAR1, CYP2D6, PLA2G4D, CYP2A6, DDX5) or quite often were simply missing from most samples. The number of CFS subjects from Perez et al. at each of these snps, and overall, is unknown. The majority of the errors with less-striking ratio inflations arose from the reported Kaviar control data where we found these to be lower frequencies than both ALFA and 23andMe control datasets for many of the snps.

The Perez et al. discussion goes through, spelling out one by one, the function of each of the genes from the top 10 in the table by summarizing what is known and including speculation for how these factors may tie into CFS. For example, they suggest that decreased metabolism of xenobiotics may be relevant to multiple chemical sensitivity disorder which is in turn relevant for some CFS cases and a gene that is downregulated by sleep deprivation which in turn is a factor in chronic fatigue states. The present reanalysis finds at the very least that such discussion and speculation are premature as there is no evidence that any of those genes are relevant.

The three genes with polymorphisms that remained with the original study criteria of occurring at least twice as often in CFS patients were EFCAB4B, MORN2 and LINC01171 which involve calcium ion channels, cell differentiation, and a long non-coding RNA transcript respectively. It is tempting to fish for connections such as to other ion channel polymorphisms that have been found relevant in CFS (8) but here too, it is premature to speculate; reanalysis of the full 23andMe data for CFS patients, as is now clearly warranted, may produce an entirely different top 50 leaderboard and functional analysis. Likewise, the criterion of a ratio of at least double may also prove too stringent which may then put some of the snps back into consideration but this too is unknown until a full reanalysis. It is beyond the scope of this article to raise that high CADD scores also may not always be an appropriate filter (e.g. 9). We conclude that the top-50 table presented in the CFS study does not reflect the top 50 deleterious differences between Chronic Fatigue Syndrome and unaffected individuals as intended but rather the top 50 errors in the 23andMe and Kaviar databases.

The present reanalysis highlights the need to use control data from the same commercial direct-to-consumer genetic testing company when used for research. On a positive note, quirks aside, there is generally high agreement between 23andMe and scientific genetic database ALFA. There are 10 million direct-to-consumer genetic test results which positively dwarfs the data collected in scientific studies. Using the abundant commercial DNA results to find genetic predispositions is very appealing especially for disorders without known cause, like Chronic Fatigue Syndrome The promise for successful continued mining of public data for research purposes remains but with caution over select snps and chips.

## Data Availability

All data referred to is provided either within the manuscript itself or supplementary materials with the manuscript.

## Author Contributions

Conceived the project (FB), extraction from openSNP (BGT), data analysis (FB, BGT), manuscript writing (FB), manuscript edit (BGT).

## Footnotes

1 https://opensnp.org

2 https://www.ncbi.nlm.nih.gov/snp

## References

1. Bedford FL. Sephardic signature in haplogroup T mitochondrial DNA. Eur J Hum Genet. 2012 Apr;20(4):441–8.

2. Perez M, Jaundoo R, Hilton K, Del Alamo A, Gemayel K, Klimas NG, et al. Genetic Predisposition for Immune System, Hormone, and Metabolic Dysfunction in Myalgic Encephalomyelitis/Chronic Fatigue Syndrome: A Pilot Study. Front Pediatr [Internet]. 2019 [cited 2020 Jul 24];7. Available from: https://www.frontiersin.org/articles/10.3389/fped.2019.00206/full

3. Cariaso M, Lennon G. SNPedia: a wiki supporting personal genome annotation, interpretation and analysis. Nucleic Acids Res. 2012 Jan 1;40(D1):D1308–12.

4. Glusman G, Caballero J, Mauldin DE, Hood L, Roach JC. Kaviar: an accessible system for testing SNV novelty. Bioinformatics. 2011 Nov 15;27(22):3216–7.

5. Tandy-Connor S, Guiltinan J, Krempely K, LaDuca H, Reineke P, Gutierrez S, et al. False-positive results released by direct-to-consumer genetic tests highlight the importance of clinical confirmation testing for appropriate patient care. Genetics in Medicine. 2018 Dec;20(12):1515–21.

6. Greshake B, Bayer PE, Rausch H, Reda J. openSNP–A Crowdsourced Web Resource for Personal Genomics. PLOS ONE. 2014 Mar 19;9(3):e89204.

7. Tryka KA, Hao L, Sturcke A, Jin Y, Wang ZY, Ziyabari L, et al. NCBI’s Database of Genotypes and Phenotypes: dbGaP. Nucleic Acids Res. 2014 Jan;42(Database issue):D975–979.

8. Marshall-Gradisnik S, Smith P, Brenu E, Nilius B, Ramos S, Staines D. Examination of Single Nucleotide Polymorphisms (SNPs) in Transient Receptor Potential (TRP) Ion Channels in Chronic Fatigue Syndrome Patients. Immunology and Immunogenetics Insights. 2015 Apr 15;7.

9. Mather CA, Mooney SD, Salipante SJ, Scroggins S, Wu D, Pritchard CC, et al. CADD score has limited clinical validity for the identification of pathogenic variants in non-coding regions in a hereditary cancer panel. Genet Med. 2016 Dec;18(12):1269–75.

