## Supplementary figures and images for "Re-analysis of genetic risks for Chronic Fatigue Syndrome from 23andMe data finds few remain"

### Supplemantal Figure 1

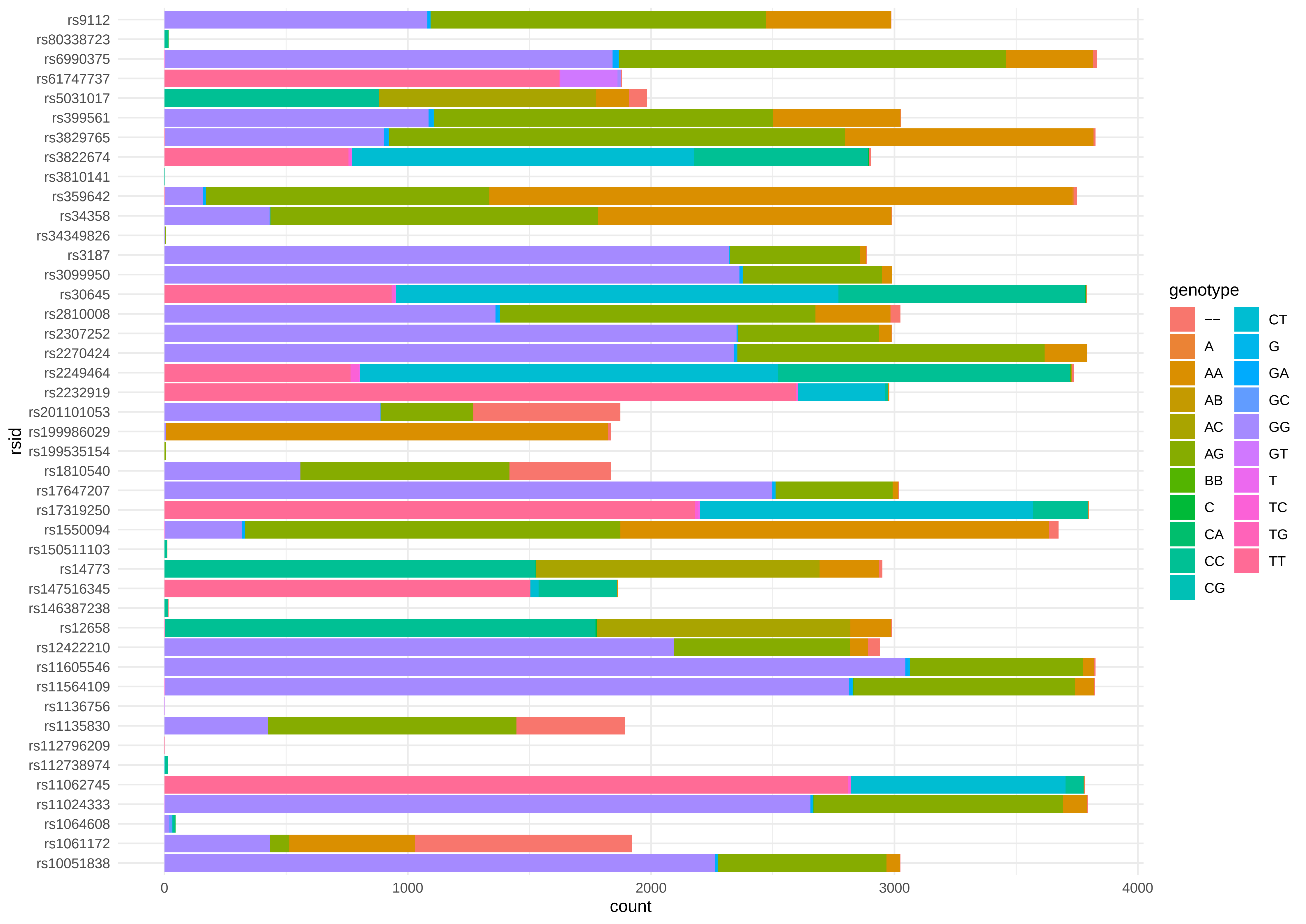
